# Identification of *Staphylococcus aureus* and prevalence of *Staphylococcus aureus* resistant to mithicillin and other patterns of resistance to antibiotics in clinical isolates of diabetic foot

**DOI:** 10.1101/2024.07.08.24310121

**Authors:** Edder Guadalupe Segura Ramon, Gabriel Martinez Gonzalez, Jorge Almeida

## Abstract

**Objective:** To determine the prevalence of methicillin-resistant *Staphylococcus aureus* (MRSA) in diabetic foot wounds and analyze antibiotic resistance patterns.

**Introduction:** The global incidence of diabetes mellitus, particularly type II, has significantly increased, leading to complications such as diabetic foot ulcers. These ulcers often become infected, with MRSA being a common and challenging pathogen. Understanding the prevalence and resistance patterns of MRSA in diabetic foot infections is crucial for effective treatment.

**Methodology:** Diabetic foot wound samples were collected from 65 patients in the Valley of Toluca, Mexico. Samples were cultured and analyzed using chromogenic agar, BHI, salt and mannitol, calf blood, EMB, and MacConkey agars. Strain identification and antibiotic sensitivity testing were performed using the Vytek automated system. Oxacillin and cefoxitin were used to detect methicillin resistance. Statistical analyses, including Kolmogorov-Smirnov, Shapiro-Wilk tests, and Spearman correlation, were conducted to evaluate relationships between clinical factors and antibiotic resistance.

**Results:** Of the 65 samples, 56.9% were from men and 43.1% from women, with 97.1% having type II diabetes. MRSA prevalence was 63%. Resistance rates were highest for ampicillin (100%), clindamycin (89%), erythromycin (87%), and gentamicin (73%). Statistical analysis showed no significant correlation between oxacillin resistance and glycemic control, erythromycin resistance (MLS resistance), hypertension, or gender.

**Conclusion:** The high prevalence of MRSA in diabetic foot wounds underscores the need for targeted infection control and appropriate antibiotic therapy. The lack of correlation between resistance and clinical factors suggests a multifactorial nature of antibiotic resistance, necessitating broader clinical and microbiological considerations for future studies.

## Introduction

The incidence of diabetes is increasing globally, reaching epidemic levels in low- and middle-income countries, which creates concern in the health care systems with limited resources and persistent challenges in treating communicable diseases.^1^ Diabetes mellitus is a major health and socioeconomic burden worldwide. Over the past few decades, the alarming increase in its incidence. The prevalence of type 2 diabetes mellitus has tripled in the past 3 decades and is expected to cross more tan 320 million by 2025.^2^

Diabetes mellitus is a heterogeneous metabolic disorder characterized by the presence of hyperglycemia due to impairment of insulin secretion, defective insulin action or both. The chronic hyperglycemia of diabetes is associated with relatively specific longterm microvascular complications affecting the eyes, kidneys and nerves, as well as an increased risk for cardiovascular disease (CVD).^3^

People with type-2 diabetes usually develop the condition after age 45, and the risk for getting it increases with age.^4^

Uncontrolled diabetes contributes to the development of neuropathy and peripheral arterial disease by complex metabolic pathways. Loss of sensation caused by peripheral neuropathy, ischaemia due to peripheral arterial disease, or a combination of these may lead to foot ulcers.^5^ Diabetic foot is a severe chronic diabetic complication that consists of lesions in the deep tissues associated with neurological disorders and peripheral vascular disease in the lower limbs. The incidence of diabetic foot has increased due to the worldwide prevalence of diabetes mellitus and the prolonged life expectancy of diabetic patients.^6^

Patients with diabetes are susceptible to infection related to immunodeficiency, neuropathy, and arteriopathy. A significant reduction in bactericidal capacity and phagocytosis may lead to dreaded complications. An infected foot ulcer accounts for ∼60% of lower extremity amputations, making infection perhaps the main proximate basis of this tragic outcome. In a large prospective study of patients with DFU, the existence o infection augmented the risk of a minor amputation by 50% compared to ulcer patients without infection.^7^

The most common microorganisms isolated from patients with DFI were reported as *Staphylococcus aureus, Pseudomonas aeruginosa, Escherichia coli, Streptococcus spp*., *Enterococcus spp*., *Proteus mirabilis* and anaerobes.^8^

Although the immunosuppression state associated with diabetes is aknown risk factor for staphylococcal infections, the influence of diabetes in the development of MRSA infection.^9^ *Staphylococcus aureus* is the most common causative agent in DFIs, and among these 23.7% were reported as Methicillin-resistant *Staphylococcus aureus* (MRSA).^10^ The most important mechanism of the propagation of MRSA and the other microorganisms is contact between people. The most common transfer is from one person to another by means of contaminated hands of healthcare/auxiliary personnel who do not wash them correctly between one patient and the next. Transmission because of contamination of healthcare material and/or surfaces has also been reported and is known as healthcare-associated infection (HAI).^11^

This pathogen presents many treatment difficulties, particularly in the provision of appropriate empiric antimicrobial therapy. Approximately 40–50% of all *S. aureus* isolates exhibit methicillin resistance which confirms almost universal beta-lactam resistance.^12^ Vancomycin, which the antibiotic that was most frequently prescribed, was given in 78% of all antibiotic regimens.^13^ Knowledge of the prevalence of colonisation or infection of diabetic patients by resistant pathogens, including MRSA, will therefore be important in assessing the extent to which interventions targeted towards diabetic patients may mitigate the spread of resistant pathogens.^14^

In the present research work, samples of diabetic foot wounds were collected to first identify *S. aureus* and the antibiotic resistance patterns shown by methicillin-sensitive strains and, on the other hand, the prevalence of MRSA.

## Methods

Diabetic foot wounds were studied in patients aged 44-years, with a mean age of -84 years, who presented different periods of time with a problem of hyperglycemia and different complications associated with diabetic foot, and exclusion criteria were not considered. With a total of - cultures, 37-belong to male patients and -28 to women. All this in the Valley of Toluca, Mexico.

To collect the sample, it was carried out under adequate hygiene conditions that would prevent cross contamination, a sterile swab was used to rub the affected area of the diabetic foot, this swab was placed in Copan transport medium, which allows the growth of aerobic and anaerobic organisms. Media was stored at room temperature until prior to primary passage for reseeding.

The primary swab was reseeded on chromogenic agar, BHI, salt and mannitol, calf blood, EMB and MacConkey, 6 hours after taking the sample. In addition to microbial reseeding, a Gram stain was performed on each of the samples. The Petri dishes were placed in an incubation oven for 18 hours at 37 ° C.

Once the reseedings were obtained between approximately 18 hours, the identification of the strain was carried out, as well as the sensitivity to antibiotics with an automated method with the VITEK^®^ 2, equipment. For the antibiotic tests tested and the concentrations, the CLSI m100 standard was followed.

To detect resistance to methicillin, the oxacillin 1 µg test and the cefoxitine 30 µg test were used. For the rest of the antibiotics, the following were tested at the indicated concentrations (Penicillin 10 µg, Ampicillin 10 µg, Ampicillin/sulbactam 8/4µg, Amoxicillin/clavulanic 10 µg, Ceftriaxone 30µg, Oxacillin 1 µg, Cefoxitin 30 µg, Clindamycin µg, Erythromycin 15 µg, Gentamicin 10 µg, Tetracycline 30 µg, Levofloxacin 5 µg, Ciprofloxacin 5 µg, Mofloxacin 5 µg, Rifampin 1 µg, Trimethoprim/sulfamethoxazole 1.25/23.75 µg, Linezolid 4 µg, Daptomycin 1 µg).

In this study, the relationship between oxacillin resistance and several clinical factors was analyzed, including glycemic control, erythromycin resistance, arterial hypertension (HTN) and the gender of the patients. To determine the normality of the data, the Kolmogorov-Smirnov and Shapiro-Wilk tests were used and the Spearman correlation coefficient was applied to evaluate the possible relationships between the variables. Statistical analyzes were performed using IBM SPSS Statistics for Windows, version 21.0 (IBM Corp., Armonk, NY).

## Results

Of 100% of the sample (n=65), 56.9% (n=37) correspond to men and 43.1% (n=28) to women, of which 97.1% (n=63) have diabetes mellitus type II and only 2.9% (n=2) type I diabetes mellitus, among the concomitant diseases, 27.7% (n=18) present additional arterial hypertension, all of them in 100 of the people with type II diabetes mellitus. For cardiovascular disease, only 16.9% (n=11) presented at least one episode, all of which corresponded to patients with DMII. The glycemic control of the patients was as follows: for people with type I diabetes mellitus, 100% had control using insulin plus an oral hypoglycemic agent; for patients with type II diabetes mellitus, only 40% (n=26) maintained control. Of the sample that are not patients with controlled DMI, only 10.7% (n=7) use insulin as glycemic control and 29.2% (n=19) have glycemic control with unspecified oral hypoglycemic agents. Only 10.7% (n=7) have had more than one reinfection, only 1.5% (n=1) of people who did not have a recurrence had MRSA. Table number 1 shows the characteristics of the patients in the study.

**Table 1:** Demographic and Clinical Characteristics of the Patients Studied. This table includes information on patients’ gender, type of diabetes, high blood pressure, cardiovascular disease, and glycemic control.

| Characteristics of the Patients | n=65 |
| --- | --- |
| Gender |  |
| Male | 56.9% (n=37) |
| Female | 43.1% (n=28) |
| Diabetes mellitus type II | 97.1% (n=63) |
| Diabetes mellitus type I | 2.9% (n=2) |
| Arterial hypertension | 27.7% (n=18) |
| Cardiovascular disease | 16.9% (n=11) |
| Glycemic control |  |
| Glycemic control DMI | 100% (n=65) |
| Glycemic control DM II | 40% (n=26) |

For the antibiotics tested, 100% (n=65) resistance to ampicillin was obtained, 86% (n=56) for ampicillin/sulbactam, which, as described below, suggests the presence of beta-actamases. In order of resistance, clindamycin showed 89% (n=58) resistance and erythromycin 87% (n=57) suggestive of MLS type resistance. For the aminoglycoside gentamicin it was 73% (n=48), tetracycline showed 67% (n=44), in the case of beta-lactams and MRSA markers; penicillin, amoxicillin/clavulanic acid, ceftriaxone and oxacillin, as well as the cefoxitin test, there is 63% (n=41) resistance. The quinolones levofloxacin and ciprofloxacin showed 56.9% (n=37) and mofloxacin 50.7% (n=33), rifampin showed only 33.8% (n=22) and trimethoprim/sulfamethoxazole 23% (n=15), for linezolid and daptomycin alone 1.5% (n=1) showed resistance. Table 2 shows the resistance patterns to the antibiotics tested for *S. aureus* and Graph No. 1 shows the patterns of resistance and sensitivity to the antibiotics tested. Where a high resistance to beta-lactams is seen, typical of resistance to methicillin in some strains and in others a production of beta-lactamases. On the other hand, it is important to highlight the high MLS-type resistance present, with the erythromycin marker. For quinolones and tetracycline there is also a high resistance of more than 50%.

**Table 2:** Antibiotic Resistance Patterns in Staphylococcus aureus Isolated from Diabetic Foot Wounds. This table summarizes the percentages of resistance and sensitivity to each antibiotic tested in the study.

| Antibiotics | Resistant | Sensitive |
| --- | --- | --- |
| Penicillin | 63% | 37% |
| Ampicillin | 100% | 0% |
| Ampicillin/sulbactam | 86% | 14% |
| Amoxicillin/clavulanic | 63% | 37% |
| Ceftriaxone | 63% | 37% |
| Oxacillin, | 63% | 37% |
| Cefoxitin | 63% | 37% |
| Clindamycin | 89% | 11% |
| Erythromycin | 87% | 13% |
| Gentamicin | 73% | 27% |
| Tetracycline | 67% | 33% |
| Levofloxacin | 56.9% | 43.1% |
| Ciprofloxacin | 56.9% | 43.1% |
| Mofloxacin | 50.7% | 49.3% |
| Rifampin | 33.8% | 66.2% |
| Trimethoprim/sulfamethoxazole | 23% | 77% |
| Linezolid | 1.5% | 98.5% |
| Daptomycin | 1.5% | 98.5% |

**Graph 1:**
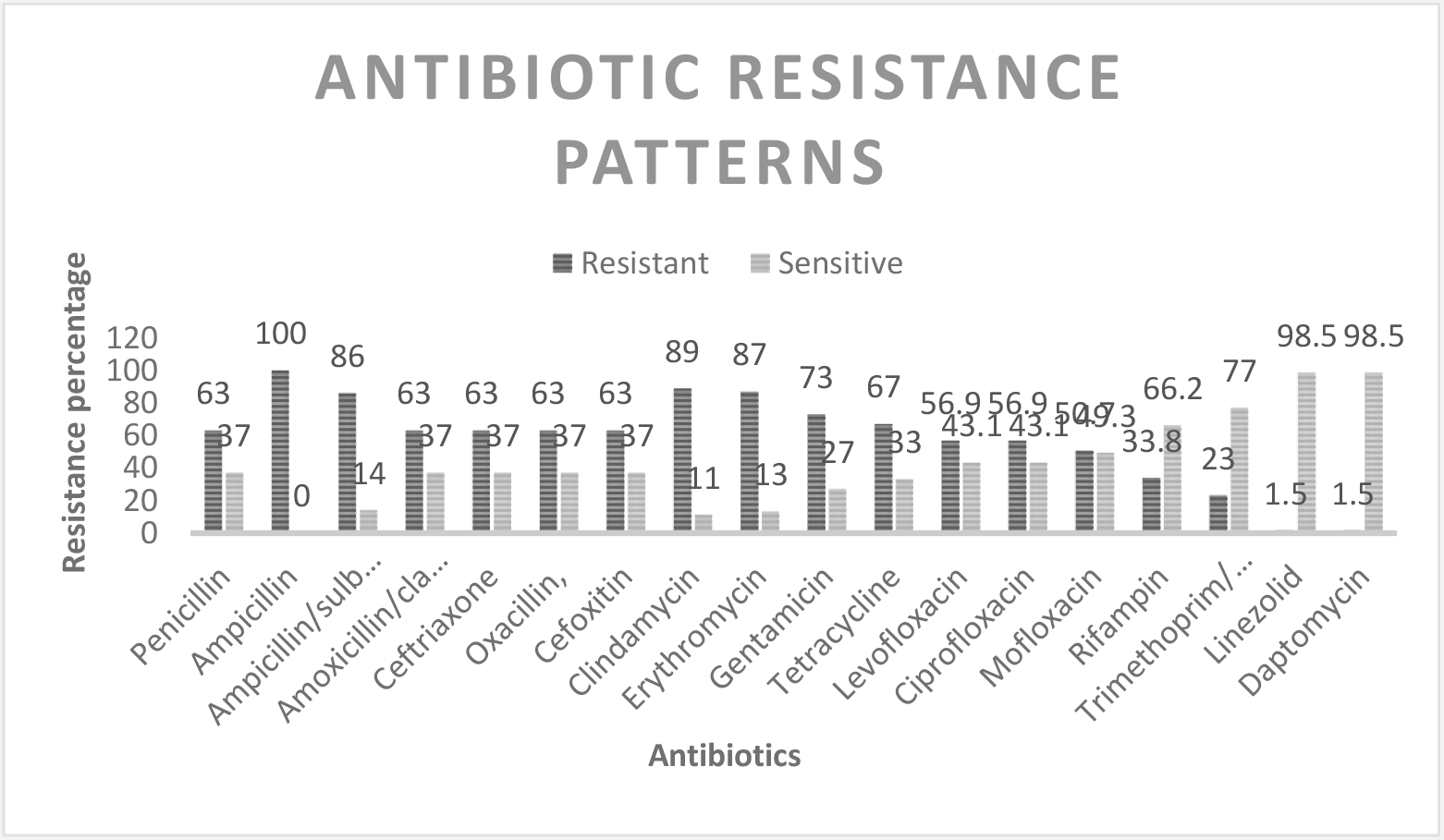
Prevalence of Antibiotic Resistance in *Staphylococcus aureus* Isolated from Diabetic Foot Wounds

## Discussion

*Staphylococcus aureus* is one of the most important causes community acquired pathogens. Antibiotic resistance is very common among strains of *S. aureus*. One of the most resistant forms of this bacterium is methicillin-resistant strains.^15^ Methicillin is used to treat bacterial infections caused by organisms of the genus *Staphylococcus*. It is active against certain types of staphylococci, which are resistant to penicillin, but ultimately show resistance to methicillin.^16^ The high prevalence of strains of *S. aureus* resistant to methicillin in the community is increasing, as shown in our study, where the population evaluated is small and yet shows 41 strains equivalent to 63% as resistant to methicillin.

Methicillin-resistant *Staphylococcus aureus* (MRSA) infection was associated with patients in hospitals and skilled nursing facilities. In recent years, reports of community-associated MRSA infections (CA-MRSA) have been increasing. Just 20 years ago in the United States, skin infections caused by methicillin-resistant *S. aureus* were observed to increase from 29% in 2001 to 64% in 2004.^17^

In a study carried out in 2014 by Lavery and collaborators in the identification of bacteria in diabetic foot, it was shown the prevalence of *Staphylococcus aureus* was 42.1%, and 70% of these isolates were methicillin resistant.^18^

In a study conducted by Lin Shin and collaborators in 2020 in Miaoli, methicillin-resistant *Staphylococcus* was found in 24.1% of the sample studied, but not a higher percentage of methicillin-sensitive *Staphylococcus*. In their study they realized that the consumption of oral hypoglycemic agents was a protective factor against *Staphylococcus* colonization in the nasal passages, but it is not described whether the same occurred in diabetic foot lesions.^9^ However, in our study, glycemic control by both oral hypoglycemic agents and insulin does not show a correlation that indicates protection against methicillin-resistant *Staphylococcus*, which is sensitive to methicillin.

In a further study carried out by 250 samples, 48 strains of S. aureus were isolated. Of which 22 presented resistance to methicillin with 45.83%, a percentage similar to that obtained in our study and which suggests the specific prevalence of resistance to methicillin in patients with diabetic foot.^19^ An important difference is that 100% of the isolates of both coagulase-positive and negative *Staphylococcus* showed 100% sensitivity to linezolid, a second-line drug when vancomycin also loses effectiveness. In our study, although it is a very low percentage, 1.5% already exists. methicillin-resistant strains with resistance also to linezolid.

In a study in Mexico in 2015 conducted by Estrella and collaborators, in the search for MRSA and its prevalence, in a sample of 100 patients diagnosed with diabetes mellitus, they found a 42% prevalence of S. aureus and of this the 34% showed resistance to methicillin.^20^ Mario Sanchez in 2017 demonstrated the prevalence of MRSA in patients with diabetic foot infection with a total S. aureus isolate of 67 strains, of which 55% showed resistance to methicillin.^21^

Another study conducted in Latin America by Gabriela Carro in 2020 showed a prevalence of S. aureus of 19% of the total isolates, of which 53.8% showed resistance to methicillin,^22^ results very similar to those we obtained in our study. The difference is that the methicillin resistance that we present is only from one region of the State of Mexico in Mexico.

On the other hand we could observe a marked MLS (macrolides, lincosamides and streptogramins) type resistance. Three mechanisms are mainly responsible for acquiring resistance to MLS antibiotics in staphylococci: (1) target site modifications by methylation or mutation; (2) active efflux of antibiotics; or (3) inactivation of antibiotics. The first mechanism includes target site modifications by a methylase encoded by one or more of the erm genes, methylating 23S rRNA and thereby altering binding sites for MLS antibiotics.^23^ In *S. aureus* is due to the action of efflux pumps, encoded by the mrsA and mrsB genes responsible for pumping macrolide and streptogramin B antibiotics out of the bacteria (MSB resistance phenotype).^24^MLSB phenotype can be expressed into forms of constitutive (cMLSB) or inducible (iMLSB).^25^

In a study conducted by Ortiz and collaborators in 2020 in a hospital in Mexico, they found 100% MLS-type resistance in S. aureus isolates.^26^

In our previous study, strains of S. aureus with an MLS-type resistance of 44% and high resistance to fluoroquinolones were identified, this in a tertiary hospital with hospital conversion during the covid 19 pandemic, in 2020.The isolated strains have the classification of MDR and XDR according to the resistance patterns shown.^27^

In the statistical análisis the results showed that there is no significant correlation between oxacillin resistance and glycemic control (Rho = -0.043, p = 0.736), erythromycin resistance (Rho = 0.102, p = 0.421), HBP (Rho = -0.096, p = 0.445) nor gender (Rho = 0.171, p = 0.172).

These findings suggest that oxacillin resistance is not influenced by glycemic control, erythromycin resistance, HTN or gender in the sample studied. The lack of significant correlation could be due to the multifactorial nature of antibiotic resistance, indicating the need to consider other clinical and microbiological factors in future studies. Furthermore, expanding the sample and including additional variables could provide a deeper understanding of the mechanisms underlying oxacillin resistance.

## Conclusion

Diabetes mellitus, especially in low- and middle-income countries, is positioned as a growing threat to global public health. Its impact on health systems is aggravated by the associated complications, among which diabetic foot stands out. This study reveals worrying data: the prevalence of methicillin-resistant *Staphylococcus aureus* (MRSA) infections in diabetic foot wounds reaches an alarming 63% in the population studied.

This scenario highlights the urgent need to implement effective measures for infection control and the adequate administration of antibiotics in diabetic patients, taking into account that first and second line medications are compromised in a high percentage. The statistical analysis of the study adds complexity to the picture: resistance to oxacillin does not depend solely on glycemic control, high blood pressure, the number of reinfections or gender. This is a multifactorial phenomenon that requires a broader approach, considering both clinical and microbiological factors.

The high resistance observed to multiple classes of antibiotics, including MLS-type resistance and fluoroquinolones, raises alarm bells. It is imperative to develop more robust strategies for the management of infections in patients with diabetes mellitus.

Our study has some limitations, such as the sample size, the lack of molecular studies to detect specific genes present in the isolated strains, among others. However, it shows a clear panorama that gives rise to more in-depth studies.

## Data Availability

All data produced in the present study are available upon reasonable request to the authors

## Acknowledgment

Pasteur Laboratorios for carrying out the laboratory tests

## Code of ethics

The present research work was accepted and approved by the ethics committee of Pasteur Laboratories On the other hand, the cultures are obtained from a routine sample, therefore, informed concentration is not necessary.

